# Defining the Temporal Progression of Normal and Nonunion Human Humeral Shaft Using Plasma Proteome

**DOI:** 10.1101/2025.09.13.25335670

**Authors:** Louis Gerstenfeld, Paul Tornetta, Serkalem Demisse, Nazanin Nafisi, Suna Chung, Trisha Chaffe, Avani Vaishnav, Ryan Kim, David Weiss, George Hanson, Joseph Patterson, Robert V. O’Toole, Paul Matuszewski, Roman M. Natoli, Renan C. Castillo, METRC

## Abstract

Current diagnostic tools are neither informative of, nor concurrent with the underlying biological processes of fracture healing. We examined if there would be protein markers in the plasma of non-operatively fixed, isolated, non-articular, humeral fractures that would be informative of these biological processes. Five follow-up visits over a six-month period were examined in 24 patients that healed (union) and 7 non-healers (nonunion). Using the first or last clinic visit as the reference, 118 proteins out of ∼7000 proteins that were assayed, showed differential expression at log^2^ fold change ≥1.3 fold and a significance of FDR ≤ 0.5. Multiple proteins related to endochondral bone development showed significant increased plasma levels during the first 12-16 weeks. Proteins associated with acute phase response, inflammation and lipid storage showed decreased levels after their initial induction. Using a time point to time point comparison for the first three follow-up visits (up to 9 weeks after injury) between healed and nonunion patients, identified 41 proteins with differential levels at p ≤ 0.05 significance, with ten overlapping the 118 proteins that tracked normal healing. The differential group associated with the nonunion patient‘s included specific extracellular matrix and acute phase proteins that showed decreased levels relative to normal healers. These data establish the progression of endochondral bone formation and place them in context to radiological and functional outcomes in human fracture healing. The identification of differentially expressed proteins in nonunion plasma suggests the potential for the development of diagnostic markers for the early intervention of nonunion.

**ClinicalTrials.gov#:** NCT05143476

**Sponsor/Funding Source:** DOD W81XWH-19-1-0796

## INTRODUCTION

Fractures are one of the most common traumas in humans and within the top five causes of work years lost to disability globally (1). The rates that extremity fractures fail to heal are highly variable and may differ from 4 to 60% dependent on the bone, the physical nature of the fracture and the patient’s comorbidities (2–9). The cost of treatment of non-unions and delayed fracture healing are significant in terms of lost worker productivity and expenses incurred for subsequent treatment (10) . Additionally, patients with a nonunion have greater overall near-term morbidity, diminished long-term health outcomes and quality of life (11–13).

Radiography (14–16), patient-reported outcomes of pain, pain interference of function, weight bearing (17) and physical examination for gross motion at the fracture site (18) are all current diagnostic tools to follow the progression of fracture healing. However, there are no clinically used biological markers for healing. Furthermore, no consensus for the diagnosis of extremity bone nonunion has been developed, and depending on the bone and individual clinical judgement, this diagnosis is made at broadly varying times from 2 to 9 months after the injury (19). While the cellular and molecular processes of fracture have been extensively studied in animals (20–22), no definitive studies have been made in humans. The studies that have been carried out in both rodents and larger quadrupeds have shown that most extremity fractures recapitulate bone development of the axial and appendicular skeleton during embryogenesis and post-natal growth and occur through the processes of endochondral ossification (20, 22).

Relative to the biology of human fracture healing, current radiological approaches and methods of physical examination do not examine any early events of stem cell recruitment or cartilage formation that occur during endochondral fracture healing. Nor are these approaches informative of any of the molecular processes that are necessary for the regenerative processes that promote fracture healing. Because there is no ability to directly assess these early biological processes of fracture healing, understanding the underlying cellular and molecular mechanisms that lead to failed fracture healing has only been inferred from studying them in animals.

A major impediment to the development of cellular or molecular markers to follow the progression of human bone healing is an inability to directly measure these processes within bone tissues as healing progresses. In this regard, tissue biopsies and body fluids are the primary tools used to assess biological processes in clinical practice. Relative to human fracture healing, the analysis of circulating T cells in the blood, the assay of microRNAs, and specific proteins in serum or plasma have been the methods that have been examined to date for their potential as biological markers for nonunion (23, 24). The analysis of proteins found in serum or plasma so far have only examined a relatively small number of proteins associated with angiogenesis, inflammation and bone turnover. The later focus on metabolic bone markers being the most prevalent proteins examined (25). The consensus that has emerged to date is that the markers commonly used to study osteoporosis: bone liver kidney alkaline phosphatase (BLK APase), collagen-telopeptide (CTX), and intact N-terminal propeptide of Type 1 procollagen (P1NP) are highly variable in their ability to detect progression of healing. However, in one recent study of 102 tibia and femur fracture patients on differing vitamin D doses, CTX or P1NP were detected at a higher level at earlier points than at later time points during healing (26).

There are two deficiencies to the blood-based approaches used to date. The first is they have focused on proteins that are associated with a limited range of biological mechanisms that might be associated with normal or failed healing. The second is that the predominant mechanisms that have been examined (angiogenesis and bone turnover) are relatively late biological events in fracture healing occurring towards the end of the endochondral process concurrent with when radiological assessments can be made. Thus, these markers do not move the diagnosis of the progression of fracture healing to an earlier window.

In the study reported here, an unbiased screen that can detect ∼7000 proteins were used to examine the human plasma proteome across the time course of normal and nonunion fracture healing. Non-operatively treated, isolated, closed, non-articular humeral fractures over a six-month course of healing were examined. We chose to analyze this type of fracture since any changes in the serum proteome would reflect either changes at the local fracture site that spilled into the blood or systemic effects that occur in response to the injury and bone fracture, and it’s healing and would not be a consequence of surgical treatment or infections that might occur. We also excluded articular injuries since such injuries could produce proteins related to cartilage repair separate from the endochondral processes of callus formation. We placed changes in the plasma proteome in context to patient comorbidities and a series of standard clinical outcomes, including radiological and pain assessments that are known to temporally track with the progression of clinical healing.

## METHODS

### Study Approval

This study was carried out under Johns Hopkins University School of Medicine (JHU SOM) single Institutional Review Board (sIRB) approved protocol administered by Major Extremity Trauma Research Consortium (METRC) Coordinating Center (MCC) at Johns Hopkins University Bloomberg School of Public Health (JHSPH). The protocol (IRB00237830) was originally approved on November 3, 2020, with current expiration date of April 20, 2026. A written informed consent approved by JHU SOM sIRB was received prior to participation.

### Data availability

Data is not available to the public, but they can be made available from the corresponding author or MCC upon request.

### Protocol Standardization

Uniform standard operating procedures were developed by consensus of the site Principal Investigators (PIs) and implemented through MCC using site training on the study protocols, procedures, and materials. After training, all sites received certification to ensure uniform clinical implementations of the study across the multiple participating sites. Compliance regarding the study protocols was monitored by local research coordinators in cooperation with the attending surgeons as consistent for all MCC studies. All study outcomes were adjudicated by the adjudication committee that were developed for this study. Outcome variables were tracked by research coordinators, evaluated, and collected in real-time through Research Electronic Data Capture (REDCap) system, and remotely monitored by the MCC research team. Site performance was assessed using weekly summary site report and monthly site coordinator meetings.

### Patient Population

Nine of the MCC-affiliated institutions across the United States participated in this study. If the site’s treating physician deemed the patient was appropriate for non-operative management, patient was recruited for the study was within 3 weeks of injury.

Patients meeting the following inclusion criteria are eligible to be recruited into the study:

1. Adult patients 18-70 years of age.
2. Isolated, closed, extra-articular proximal humerus metaphyseal fractures or humeral shaft fractures (Arbeitsgemeinschaft für Osteosynthesefragen/Orthopaedic Trauma Association (AO/OTA) classification system11 type A2.1, A2.2 and/or A2.3) with diaphysis involvement or diagnosis of isolated closed extra-articular fractures of the humeral shaft (AO/OTA 12 A, B, or C) .
3. Consent for non-operative management of their fracture.

Any patient meeting one or more of the following criteria were excluded from the study:

1. Articular involvement
2. Open fractures
3. Additional long bone injuries
4. Injury not amenable to non-operative management
5. Pregnancy
6. Immuno-compromised state

Our study examined male and female humans, and similar findings are reported for both sexes.

A total of 377 patients were screened across 8 sites with a total of 41 patients enrolled for 24 months. Thirty-one of these patients had sufficient data and blood draws for analysis, with 24 assessed as healed and 7 assessed as nonunion. Supplemental table one provides a complete breakdown of patient demographics comorbidities and primary and secondary data that were collected from the final cohort of patients used for this study.

### Outcome Variables

Table one in supplementary document section summarizes the clinical information and outcomes that were collected.

Primary variables included:

1. modified Radiographic Union Score for Tibia (mRUST)
2. Patient Reported Outcomes Measurement Scores (PROMIS):

a. Social Participation
b. Physical Function
c. Pain Intensity
d. Pain Interference
e. Veterans RAND 12 Item Health Survey (VR-12) Mental and Physical Component Scores (VR12 MCS and VR12 PCS).

At each study visit clinical and radiographic assessments of healing by the treating physician, blood draws for plasma protein analysis, and administration of patient-reported outcome measures were performed.

### Plasma Protein Analysis

Blood samples were collected in standard anti-coagulant vacuum tubes, placed on ice and processed within 30 minutes of the drawing. All whole bloods were centrifuged to isolate the plasma, aliquoted and froze at -80 Celsius within 2 hours of the blood draw. Samples were thawed once, and final aliquots of 200 microliters were made and shipped on dry ice to SomaLogic Inc, a private protein biomarker discovery and clinical diagnostics company. Proteomic analysis used the value normalization of ∼7000 proteins screened by SomaLogic Inc, Aptamer platform.

### Statistical Analysis

Two main statistical analyses were conducted. Prior to analyses, patients were classified into union (normal healing) and nonunion groups using two sets of healing criteria. The stringent (S) criteria for normal healers required both a mRUST score greater than 12 and the absence of motion at the fracture site to confirm healing. In contrast, the laxer (L) criteria relied on clinical examination findings consistent with healing, which included cases where either the mRUST score was below 12 or the radiographic findings were inconclusive. S criteria used to define nonunion was made at the third recall period (7-9 weeks). It was based on having an mRUST score of 6 or less and having motion at the site of fracture on physical examination. A non-stringent assessment was based only on clinical examination. All protein measurements were log2 transformed prior to all analysis.

1. Clinical analysis: Continuous variables were summarized using means and standard deviations, and categorical variables were summarized using counts and percentages. Comparison of clinical variables between patients who healed their fractures and those that had non-unions was performed using Student’s t-test and Mann-Whitney for normally and non-normally distributed continuous variables respectively, and Chi-square test or Fisher’s exact test for categorical variables.
2. Proteomic Time Course Assessments: Initially, for union patients with normal healing outcome, temporal changes in protein levels were assessed using mixed-effect linear regression models (Proc MIXED, SAS), where the proteomes (as dependent variables) were modeled as a function of time (time/visit considered as a categorical independent variable). The primary comparison focused on identifying protein changes from the initial (baseline) visit, whereas a secondary evaluation assessed differences from the final time point. Proteins with a fold change threshold of ≥1.3 and a false discovery rate (FDR) <0.05 were considered statistically significant and identified as differentially expressed proteins (DEPs). The mixed-effect linear regression model used here is similar to a paired-t test but utilizes data more efficiently by examining all time points within a single model while accounting for correlated data. However, due to limited data that prevents the consideration of more complex correlation structures, we employed only an exchangeable correlation structure.
3. Analysis of Proteins in relation to Union/Nonunion outcome: In the second analysis, nonunion patients were compared to union patients, focusing on the initial three post-injury time points to identify early molecular differences that may serve as potential diagnostic biomarkers before radiographic detection. For this analysis, only the first three recall visits (V1, V2 and V3) and the average of the first three recall visits (M) were examined. We chose to limit the analysis to these initial recall times, to define early biomarker differences between healed and non-union patients up to the time of radiographic confirmation. Due to the limited number of nonunion cases, a less stringent threshold was applied to ensure sensitivity in detecting potential biomarkers. Proteins were considered differentially expressed if they met the significance criteria of p ≤ 0.05 (uncorrected p-value) and a fold change ≥ 1.3.

### Bioinformatics Analysis

Statistically enriched terms, including Gene Ontology (GO) and KEGG pathways, canonical signaling pathways, and hallmark gene sets, were identified using Metascape (27) based on accumulative hypergeometric p-values and enrichment factors applied as filtering criteria. To identify upstream regulators, the Ingenuity Pathway Analysis (IPA) comparison was performed, and the top 10 upstream regulators were selected based on their z-score values. To compare omics changes across species, we analyzed our human data in conjunction with previously published mouse data (28). Overlapping proteins between the human and mouse proteomic datasets were first identified, and selected proteins were then used to compare transcriptomic and proteomic expression in mice with proteomic expression in humans.

### Visualization

Volcano plots and heatmaps were generated using the pheatmap package in R, while whisker plots were created using GraphPad Prism version 10.

## Results

### Assessment of Clinical Progression of Healing

A breakdown of the primary demographic and clinical comorbidities associated with the patients enrolled for this study are summarized in Table 1 with a more extended summary in the supplemental data. The primary features that showed the strongest association to healing were the mRUST scores and the PROMIS Pain scores. A summary of their significance is seen in Table 2 while a graphical association of how these patient-related clinical features change over time is in Figure 1. Patients determined as nonunion at 7-9 weeks had lower mRUST scores at every recall time even prior to 7-9 weeks when the nonunion determination was made. They all failed to show any increase over time in contrast to the healed group that all showed a continued increase in mRUST up to the last recall period. Relative to pain, all patients showed diminishing levels of pain intensity and pain interference as a function of time, however, those patients assessed as nonunion had higher pain intensity and pain interference with functional activity at all times. It is interesting to note that while the PROMIS for pain intensity showed significance at only the 4–6-week period, that patients reported functional pain interference for up to 12 weeks.

**Table 1.** Demographics and Injury Characteristics.

|  | **Entire Cohort** | **Union** | **Nonunion** | **p-value** |
| --- | --- | --- | --- | --- |
| **Age (years)** | 42.25 ± 16.88 | 41.24 ± 17.88 | 49.50 ± 11.50 | 0.39 |
| **Sex** |  |  |  | 1.000 |
| *Male* | 10 (31.3%) | 7 (41.2%) | 1 (25%) |  |
| *Female* | 22 (68.8%) | 10 (58.8%) | 3 (74%) |  |
| **Injury mechanism** |  |  |  | 0.03 |
| *Low energy* | 13 (40.6%) | 6 (35.3%) | 2 (50%) |  |
| *Moderate energy* | 13 (40.6%) | 10 (58.8%) | 0 (0%) |  |
| *High energy* | 6 (18.8%) | 1 (5.9%) | 2 (50%) |  |
| **Fracture pattern** |  |  |  | 1.000 |
| *One fracture line, two pieces* | 15 (46.9%) | 8 (47.1%) | 2 (50%) |  |
| *More than one fracture line, multiple pieces* | 17 (53.1%) | 9 (52.9%) | 2 (50%) |  |

**Table 2:**
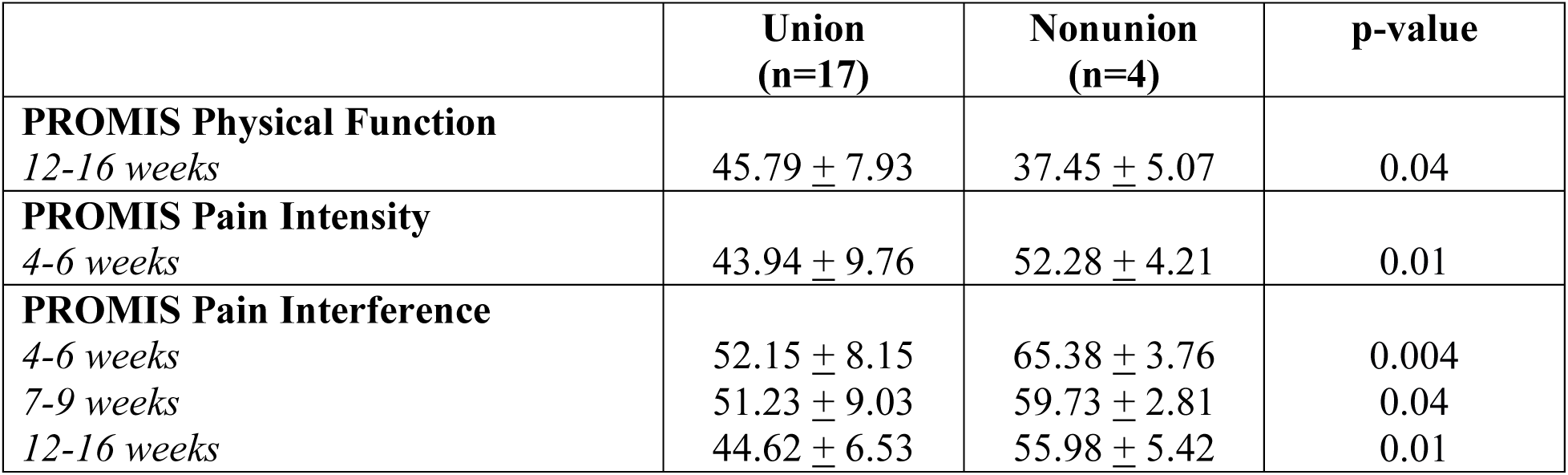
Statistically Significant Patient-Reported Outcome Measures Between Union and Nonunion Patients.

**Figure 1.**
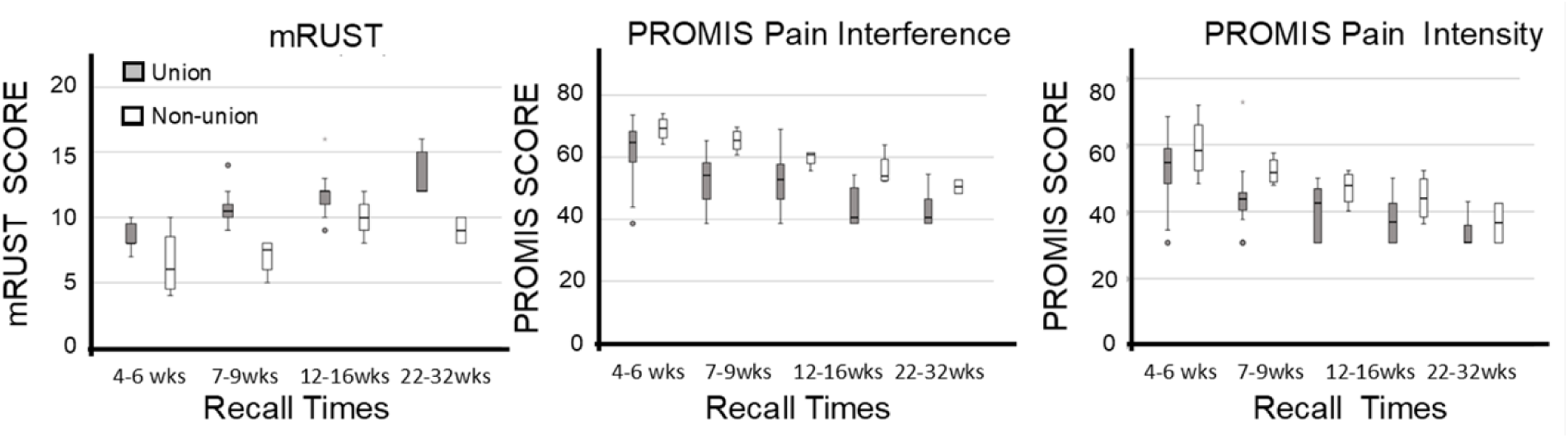
Clinical Assessments of Fracture Healing. Left Panel depicts mRUST scores, middle Panel depicts the PROMIS score for Pain Interference, while the Right most graph depicts the PROMIS scores for pain intensity. All graphs are against recall time. Bars depict the range, while the heavy line in each whisker bar is the mean. Outliers of greater than 1 Std Dev are denoted with an asterisk.

### Proteomic Analysis

Plasma proteomics analysis was conducted in two parts: (1) assessment of temporal changes in the union group, identifying significant proteins with fold change ≥1.3 and FDR ≤0.05; and (2) comparison of union and non-union groups at the same time-points, using fold change ≥1.3 and p-value ≤0.05 as significance criteria (29).

Two groups of DEPs were compared. One group was based on using a stringent criterion of healing (S) where both mRUST greater than 12 and no motion at the site was used to define if the patients had healed. The second DEP group used a laxer criteria (L) that met the clinical examination criteria, but the patient did not achieve an mRUST of ≥12 or the X-rays were indeterminate or unavailable. Figures 2A and 2B present volcano plots comparing the overall spread of the numeric values for the protein expression in union patients at the first visit, using the laxer (L) and stringent (S) healing criteria, respectively. As can be seen, the range of FDRs for several proteins were in excess to log10-12 and FC log2 x 2.5. In the L group, 68 proteins were upregulated and 50 were downregulated, whereas the S group showed 40 upregulated and 29 downregulated proteins. As anticipated, the more stringent criteria yielded a smaller set of DEPs (69 total), compared to 118 identified under the L criteria. Table 3 presents the numeric summary of this data as well as for our analysis of the union and nonunion DEPs that is presented below. The complete data sets for these comparisons using both first and last recall times by the FC, FDR and p-values for comparisons to both references are included in supplemental data for the study.

**Figure 2.**
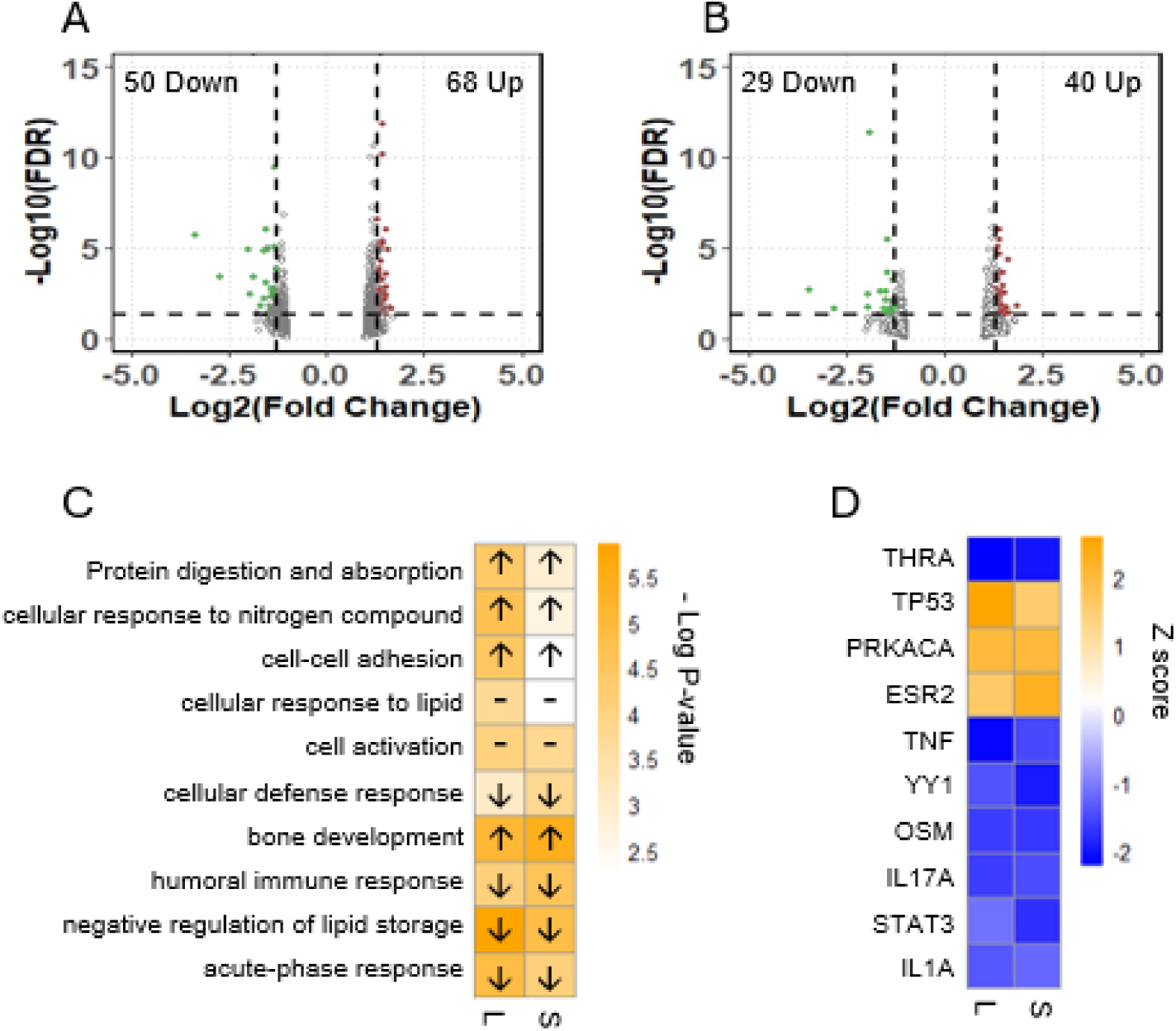
Biological Regulatory Mechanisms Associated with the Plasma Proteome of Human Fracture Healing. A) Volcano plots of the two DEP groups for the laxer (L) with FDR ≤ 0.05 and log2 FC ≥ 1.3, demonstrating 50 downregulated DEPs and 68 upregulated DEPs. B) Volcano plot for the stringent group (S) with 29 downregulated DEPs and 40 upregulated DEPs. C) Top 10 gene ontology terms for both groups based on the p-value. Upward and downward arrows indicate activation and inhibition of the terms respectively. The dashed line indicates the undecided direction of the pathway. D)Top 10 identified upstream regulators between groups based on Z score.

**Table 3.**
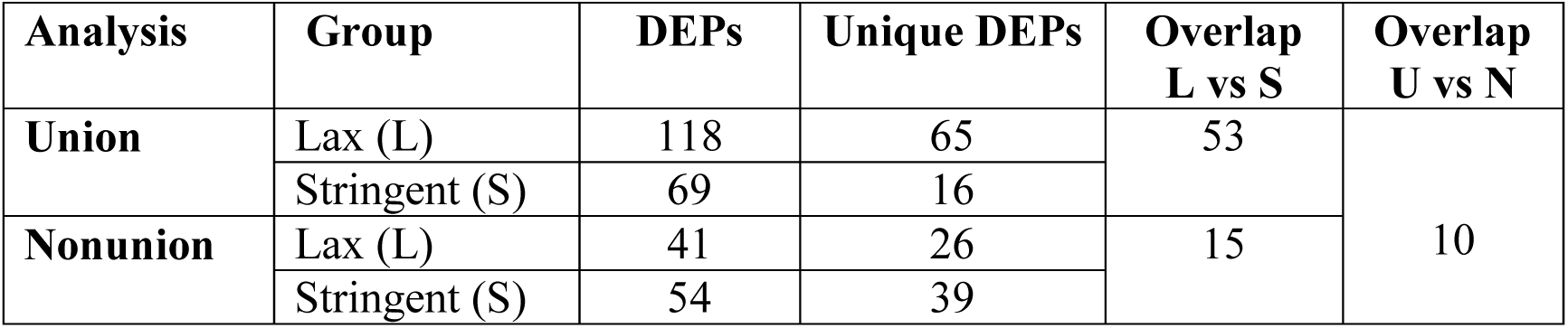
Number of Differentially Expressed Proteins (DEPs) by Overlap Between Groups.

We analyzed the biological ontologies and the upstream molecular mediators that could be inferred from the DEPs. Figure 2C displays the top 10 biological ontologies of the DEPs in both the L and S groups, along with their associated p-values. Arrows within each cell indicate the predicted direction of ontology regulation, upward for activation and downward for inhibition, while dashed lines represent ontologies with undetermined directionality. Among the top five groups, proteins related to bone development showed an increased expression while those associated with acute phase response, inflammation and lipid storage all showed a decreased expression. Figure 2D shows the top 10 predicted upstream molecular regulators associated with the DEPs in both groups. Orange cells represent activated regulators, while blue cells indicate inhibited regulators.

Figure 3 displays expression profiles for eight representative proteins from the stringent DEP group, four involved in cartilage development (grey bar) and four associated with the acute phase response and inflammation (striped bar). Time points at which individual protein levels reached the criteria for statistical significance are indicated within the figure. Asterisks (*) and double asterisks (**) indicate proteins that met the selection criteria of log2 FC ≥1.3 and FDR ≤0.05 when using either the first or last visit as reference points, respectively. Notably, Col2A1 exhibited the earliest and highest expression, followed by Col9A1 and Col10A1, which overlapped in significance during the 4-to 9-week period. COMP peaked later, between weeks 7 and 16. In contrast, markers of the acute phase response showed a pronounced elevation within the first three weeks post-injury, followed by sustained suppression over the subsequent months. In the next set of analysis, we compared the patient group that reached union to those that were diagnosed with nonunion. Using the S criteria to define our groups for comparison identified 54 differentially expressed proteins, while the L criteria yielded 41 (Table 3). The resulting top 10 biological ontology terms for DEPs are shown in Figure 4A and B, based on the S and L healing criteria, respectively. Arrows within each cell indicate the predicted direction of ontology regulation of the union samples relative to non-union. Ten DEPs were found that they overlapped between both union and nonunion groups (Table 3), and their temporal expression profiles are illustrated in Figure 5. These proteins were selected based on their significant differences in plasma levels between healed and nonunion patients (p ≤0.05 and |log2FC| ≥1.3), as well as their relevance to normal healing dynamics. All proteins meeting these criteria are shown in Figure. Proteins involved in cartilage formation or extracellular matrix (ECM) remodeling are displayed at the top, while those related to immune and acute phase responses are shown at the bottom. Apart from TREM2 and CDH17, all proteins exhibited reduced expression in nonunion patients relative to those that healed.

**Figure 3.**
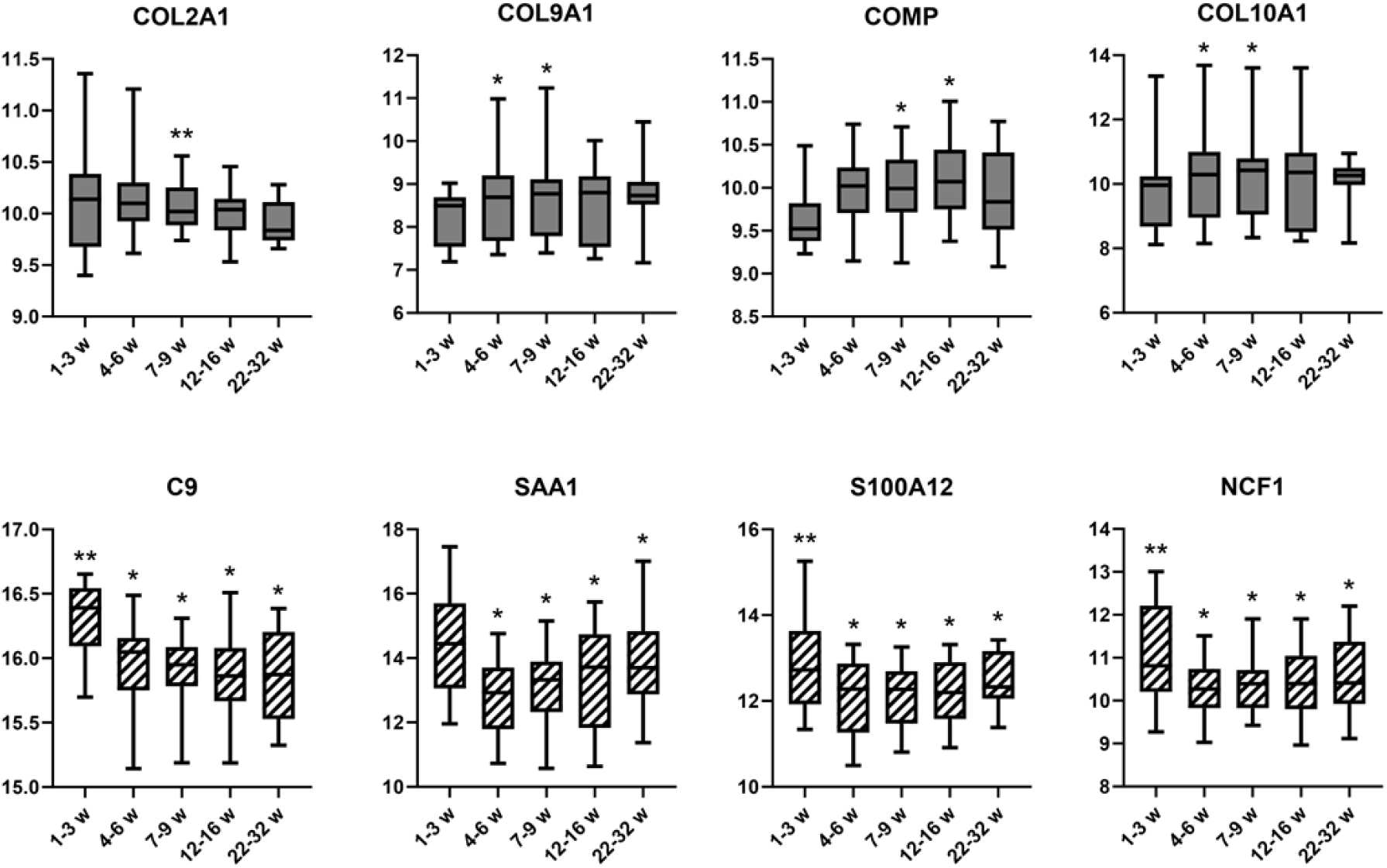
Individual Proteins within the Plasma Proteome for DEPs of Normal Healers Representative of Bone Development and Acute Phase Response. Whisker plot of proteins related to: (A) cartilage formation grey bars; and (B) acute phase (cross hatched bars). Levels over time for union samples. Asterisks (*) and double asterisks (**) indicate proteins that met the selection criteria of log2 FC ≥ 1.3 and FDR ≤ 0.05 using first or last recall visit as reference points, respectively. Mean values per protein are denoted with the heavy line.

**Figure 4.**
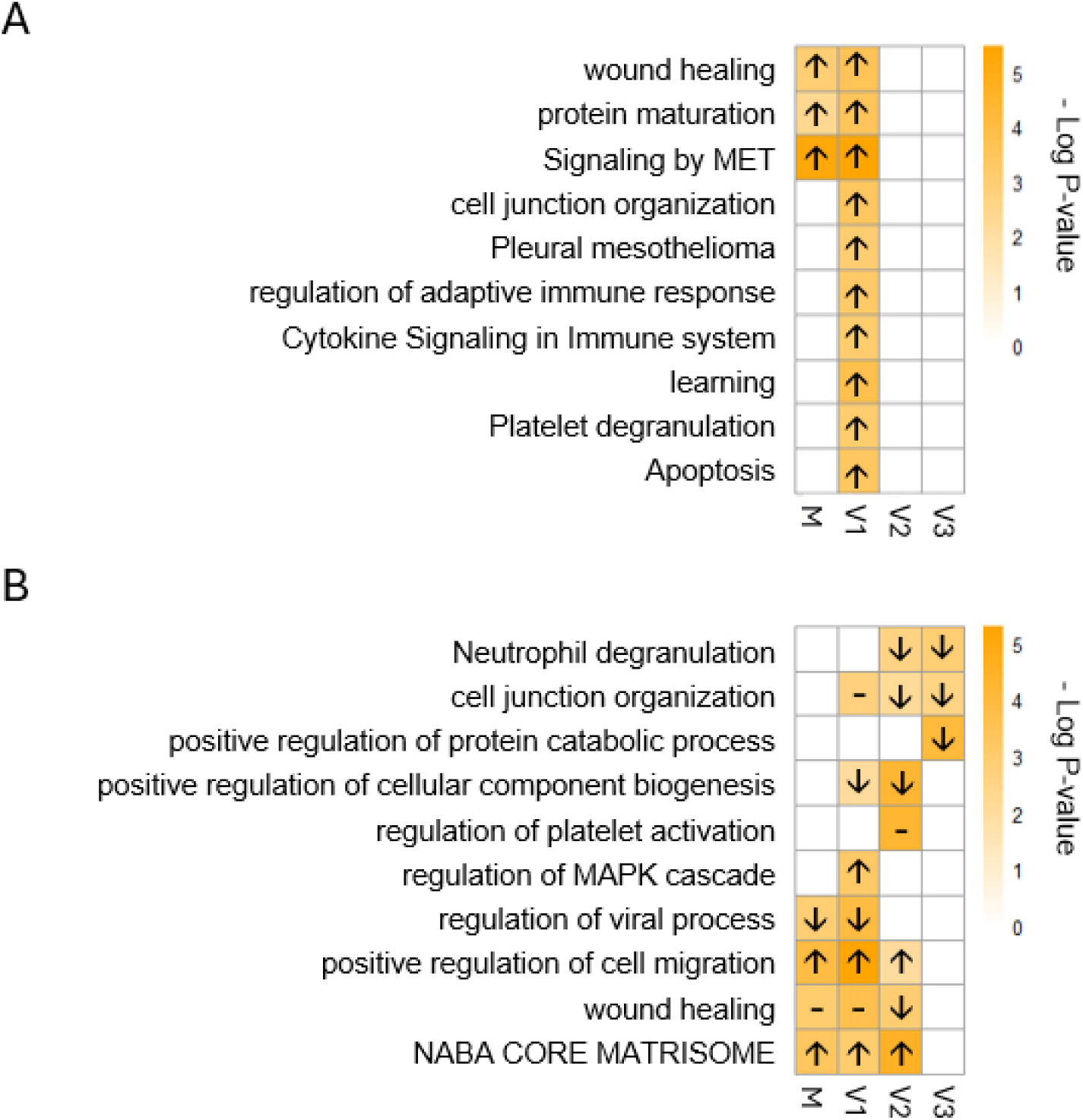
Biological Ontologies Associated with the Difference in Plasma Proteome of Nonunion Human Fracture Healing. A and B) Top 10 enriched Gene Ontology terms in the non-union group compared to the union group for: (A) Stringent; and (B) Laxer assessments, ranked by p-value. Analyses were conducted across the Mean group (M), as well as individual time points corresponding to the first (V1), second (V2), and third (V3) clinical visits.

**Figure 5.**
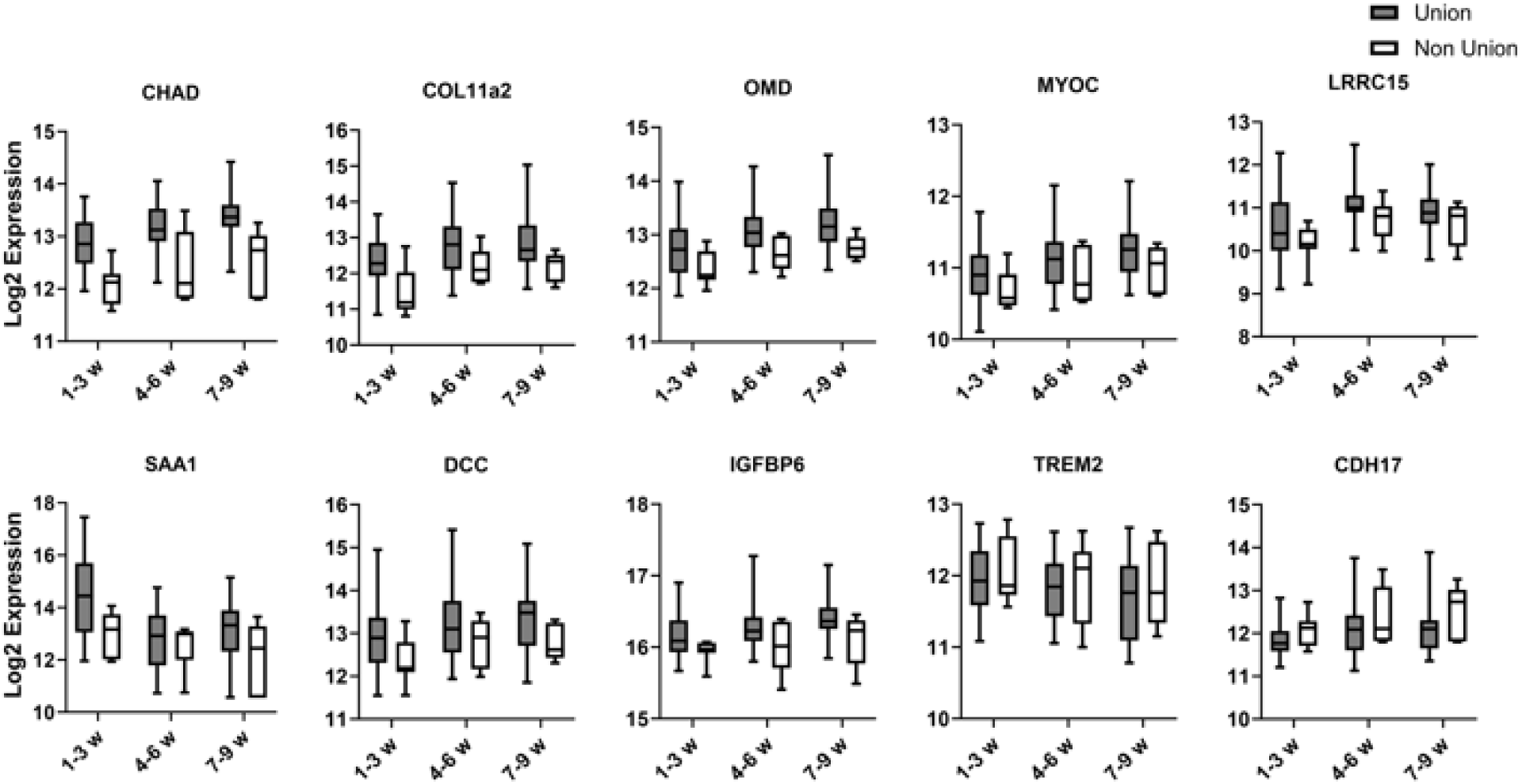
Selected Protein Difference in Plasma Proteome of Nonunion Human Fracture Healing. A and B. Temporal expression profile of overlapped DEPs between union and nonunion analysis. Gray boxes represent data from union patients, while white boxes correspond to nonunion patients. Error Bar shows the spread of 1 Std Dev, and the heavy line in each bar is the mean for the protein levels.

In the last part of this study, the human findings were compared to prior transcriptomic and proteomic studies in mice (30). Due to the limited coverage of the earlier mouse study-which utilized a smaller SomaLogic platform (∼1,300 proteins compared to ∼7,000 in this human study)-many cartilage-associated proteins identified in the human DEP dataset were not represented in the mouse proteomic analysis. Despite this, 17 DE proteins were shared between the human and mouse datasets. Figure 6 presents four representative proteins (OMD, ENG, PTN, THBS4) selected for cross-species comparison, illustrating their expression patterns in mouse transcriptomic data (Panel A), mouse proteomic data (Panel B), and human proteomic data (Panel C). In mice, the temporal patterns of mRNA expression in the fracture callus closely align with the corresponding protein levels in plasma. Although the human protein expression profiles exhibit similar overall trends to those observed in mice, the timing of these changes differ substantially between species.

**Figure 6.**
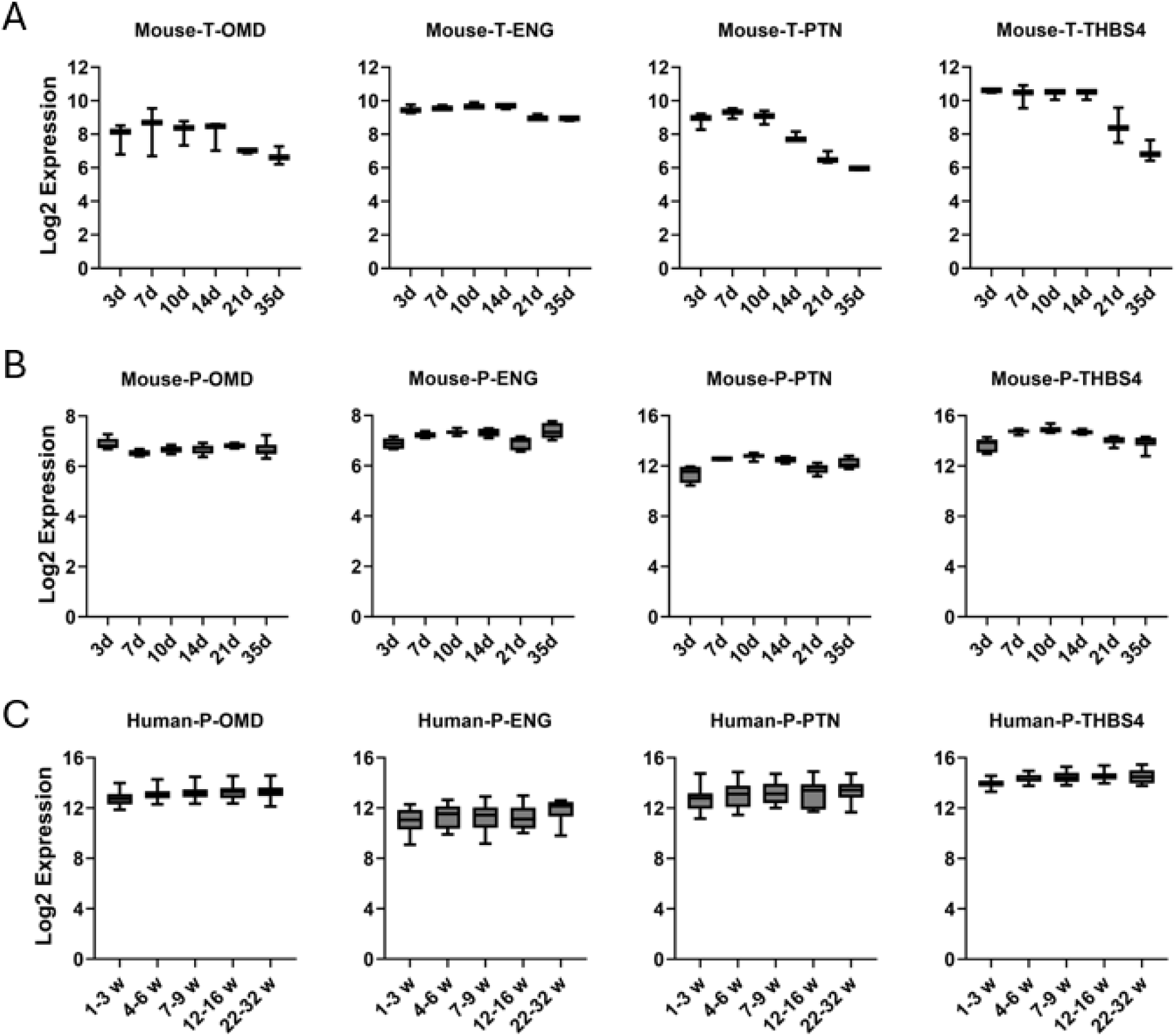
Cross-specie Comparison of the Callus Tissue mRNA and Plasma Temporal Expression of Four Proteins in Mice Relative to Their Plasma Expression in Human. (A) Relative mouse callus mRNA expression; (B) Relative mouse plasma protein level; (C) Relative human plasma protein levels

## Discussion

### Assessment of Normal Fracture Healing

We assessed how patient plasma proteomes might be informative of either local or systemic biological processes that were affected by an extremity fracture. The multiple proteins unique to both chondrocytes and the synthesis of connective tissue matrices that had differential levels of expression in the plasma may be inferred to be representative of the regenerative processes of the injured bone. Three observations are supportive of this conclusion.

1. The temporal patterns of the specific cartilage associated proteins that were observed in the plasma follows their known temporal molecular and immunohistological patterns of expression seen in developmental studies of endochondral bone formation and studies of fracture healing in animal models (30, 31).
2. The appearance of COL10A1 as a marker of endochondral bone formation of extremity bone healing is consistent with studies of tibia fractures in mice (32). In this study it was shown that this protein was seen to peak in the blood concurrent with its peak mRNA expression in fracture callus (32). In a subsequent study of human tibia and femur fracture healing, differential COL10A1 levels in plasma were also observed during the time course of healing (33).
3. Comparative analysis of our own prior murine studies (28) with our human data identified that multiple connective proteins appear in plasma after fracture that are temporally concurrent with their mRNA expression within the callus tissues. A rough extrapolation of the temporal scales of fracture healing between our mouse and human studies suggests that one day in the mouse correlates to about 1 week of human healing. Using this rough scale, the endochondral period in human humeral fracture healing is confined to 12-16 weeks post injury, while in tibia or femur fracture healing in mice the endochondral stage occurs between 12 and 18 days. Consistent with our extrapolation of the comparative temporal scales between mice and humans based on biological markers, studies comparing mRUST radiological scoring between the two species showed that the first three weeks encompassed the hematoma phase while a period between 22-65 days encompassed the endochondral period (34). The comparisons between lower and upper extremities would suggest the endochondral developmental program within different long bones in humans has a relatively constant period in which it occurs, while the later periods of coupled remodeling may be much more variable (34).

While the data for connective tissue and bone-associated proteins likely reflect local effects occurring in the callus, it is more difficult to infer the tissue origins for other proteins showing differential levels in plasma. Since many of these proteins are associated with immune cells and inflammatory responses, they could be derived locally from either cells at the fracture site or could arise from immune cell populations that are activated in multiple tissues including the spleen, marrow, or surrounding lymph nodes. The liver however is the most likely source for several specific proteins associated with the acute phase response, including serum amyloid A1, NCF1 C9, and HAMP. All the acute phase proteins that were observed in the DEP group are known to be predominantly synthesized in liver and expressed as a response to trauma (35, 36). Consistent with the idea that the liver plays an important role in injury response to bone fracture are a number of studies that suggest cross talk between the liver and bone (37, 38). More specifically, the liver has been recently shown to undergo immune cell population changes and release of immune and bone modulatory factors in response to bone fracture (39).

A primary goal of this study was to use the DEP of the human plasma proteome to gain insight into the underlying biological cellular and molecular processes that are associated with the progression of normal and failed human fracture healing. Other than bone development, the primary ontologies associated with the changes in the proteome were related to immune responses and lipid metabolisms. It is important to note that because the downregulated DEPs were made relative to the first assay period, they would all be highly elevated relative to the first recall time used as the reference. Therefore, in this context, the predicted upstream regulators for these proteins seen in Figure 2D would be highly elevated in the first three weeks after fracture even though they are indicated as being suppressed. It is noteworthy that all these regulators (OSM, TNF, Il17A, IL17a, and STAT3) control aspects of immunity and inflammation, which would be consistent with our prior animal studies showing that all these regulatory proteins were highly elevated immediately after injury (40). The inferred activity of STAT3 and IL17a is noteworthy since these proteins are associated with the control of Treg-cells and can be targeted by many biological and pharmacological agents. This also has relevance to fracture healing since Treg cells have been suggested to play an important stimulatory and protective role for skeletal stem cells and cartilage tissues during the early periods of fracture healing (40–42) in animal models (43). The importance of Treg cell function in fracture healing is also strongly supported by studies that have shown that their dysregulation in both animal and humans’ studies is associated with delayed or failed fracture healing (44).

### Assessment of the Biology of Nonunion

In our analysis of the DEP groups for both normal healing and in the comparison between healed and nonunion patient groups, we used either a more S or L criteria for the selection of the patient groups. This has relevance to the potential use of proteomic markers for diagnostic assessments since the same ontologies and upstream regulators were identified even when L criteria were used to identify if a patient is healing. However, in our analysis of nonunion patients, the S classification captured the more consistent expression differences in the repeat measurements across the three time-points, likely reflecting a stronger molecular distinction in patients with a more definitive healing outcome compared to those with indeterminate clinical assessments. This was previously noted in assessing the predictive value of regression models in epidemiological studies that attempt to estimate the ‘functional’ relationship with one or more ‘underlying’ predictor variable, which showed that regression slope between a response and predictor variable are underestimated when the predictor variable is measured imprecisely (45). This is also consistent with the observation that those patients identified at 7-9 weeks as nonunion retrospectively had the lowest mRUST in all their prior clinic visits.

Using the comparative DEP data to inform on the biological characteristics identified some overlapping ontology features related to the ECM production and immune and inflammatory activities between the healing and nonunion groups. Contrary to our expectation though, many of the individual proteins identified were unique, although given the small sample size of the nonunion group and L criteria of statistical selection, we limited the specific identification of potential markers of nonunion to the subset that overlapped with the more stringently selected healer group. It is interesting to note that all ten proteins showing overlap demonstrated differential levels of expression at the first recall period (1-3 weeks after injury) and continued to show altered levels of expression up to the time of definitive nonunion determination at 7-9 weeks. This observation would suggest that the conditions that lead to nonunion are emergent at a very early time in the healing process. Also, contrary to our expectation was our observations that many of the major extracellular matrix proteins associated with endochondral cartilage formation such as collagen types nine or ten or aggrecan did not show significant changes in the plasma of nonunion patients. The one exception was collagen type eleven which was significantly lower along with several matrix adhesions proteins and proteins that promote matrix assembly. Such results suggest that the primary problems associated with nonunion might not be the progression of chondrocyte differentiation but specific functions that promote their interactions with the matrix and their ability to assemble the matrix. Another surprising finding was that the other group of proteins that showed overlap and differential regulation in nonunion were all associated with the acute phase response.

While no comparable studies have used the SomaLogic platform to survey the proteome in patients with fracture, two recent studies were carried out using mass-spectroscopy (MS) analysis comparing normally healing human fractures and nonunion (46) or nonunion patients to healthy controls (47). In comparing the results from fractured patients to nonunion patients two proteins Saa1 and AZGP1 (data not shown) were observed in our nonunion DEP group that were identified in the (MS) analysis to have differential levels in serum (46). However, in this study the comparisons showed that Saa1 increased in the nonunion group, and the study compared fractures at about two weeks after injury to nonunion diagnosed at about a year. The other feature to note upon from this study was that they also carried out a metabolomic comparison between the two groups and identified that lipid metabolism appeared to be dysregulated in the nonunion patient group. Comparing our results with the other MS study identified multiple acute phase proteins that matched the proteins seen in normal healing in our study. In this study though the researchers compared nonunion patients to healthy unfractured controls (47). Their observations compared with our findings would then suggest that the acute phase induction remained unresolved in nonunion patients.

In summary, this study is the first use of a large-scale plasma proteomic assessment to interrogate the biological progression of human fracture healing. Multiple cartilage and extracellular matrix associated proteins were observable in the plasma that enabled temporal evaluation. These data place the endochondral phase of bone formation within the first three to four months after injury. While limited in number, the analysis of nonunion suggested that variations in plasma biomarkers may be used to identify patients who will heal versus not at very early time points (one–two months after the initial injury) compared to current standards that wait 6 or more months to declare nonunion.

## Supporting information

supplemental files for Defining the Temporal Progression of Normal and Nonunion Human Humeral Shaft Using Plasma Proteome

## Data Availability

All data produced in the present study are available upon reasonable request to the authors

## Author Contributions

LG contributed to the study design, data interpretation, writing, providing resources, and research oversight.

PT III contributed to the study design and writing.

SD contributed to statistical analysis plan, data analysis, and writing.

NN contributed data analysis and writing.

SC contributed to the study design, research oversight, writing, and providing resources.

TC contributed to the study design, research oversight, writing, and providing resources.

AV contributed to data and sample collection, data analysis, and writing.

RK contributed to data and sample collection and writing.

DW contributed to the study design, writing, and providing resources.

GH contributed to the study design, writing, and providing resources.

JP contributed to the study design, writing, and providing resources.

RO contributed to the study design, writing, and providing resources.

PM contributed to the study design, writing, and providing resources.

RN contributed to the study design, writing, and providing resources.

RC contributed to the study design, data interpretation, writing, providing resources, and research oversight.

## FUNDING SUPPORT

### Name of the Funder

Department of Defense (DoD)

### Grant Number

W81XWH-19-1-0796

### Recipient Author

Louis Gerstenfeld, PhD

## ACKNOWLEDGEMENT

We appreciate the participating sites for dedicated work on patient enrollment and data and sample collection that made this research possible.

## SUPPLEMENTAL MATERIALS

**Figure 1.**
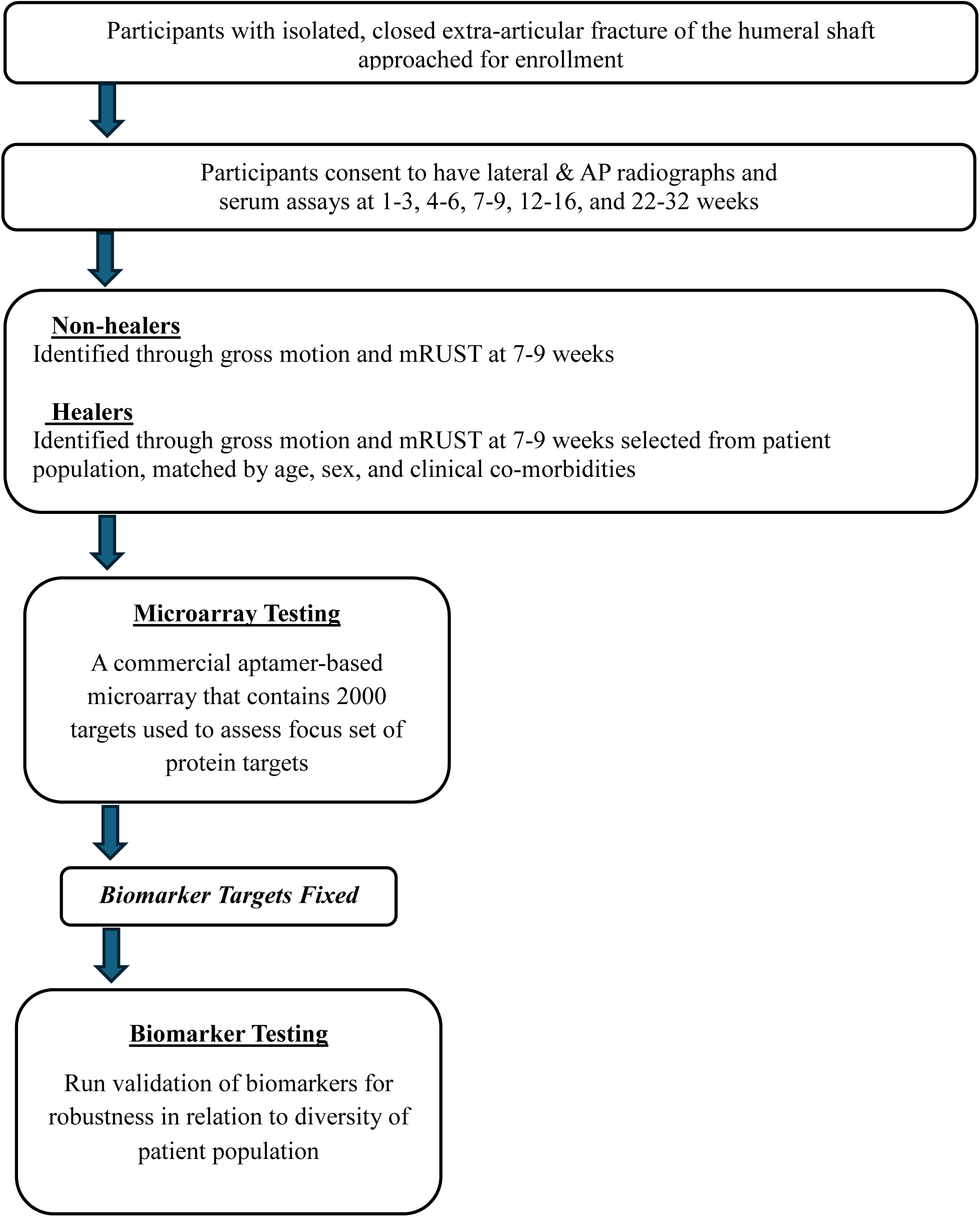
Study Design.

**Table 1.** Data Collection and Follow-up Timeline.

| **Data Collection** | **Baseline**<br>(1-3 weeks<br>since study injury) | **Follow-up**<br>4-6 weeks<br>7-9 weeks<br>12-16 weeks<br>22-32 weeks | **Diagnosis of**<br>**Nonunion**<br>(7-9 weeks) |
| --- | --- | --- | --- |
| **Inclusion/Exclusion** | X |  |  |
| **Informed Consent** | X |  |  |
| **Patient Characteristics** | X |  |  |
| **Medical History** (*Including height, weight, comorbidities and tobacco use*) | X |  |  |
| **General Injury Characteristics** | X |  |  |
| **Study Injury Characteristics** | X |  |  |
| **Hospitalization and Surgery** | X |  |  |
| **X-rays of Study Injury** | X | X |  |
| **Clinical Assessment** (*including range of motion*) | X | X |  |
| **mRUST** | X | X |  |
| **Hospital Admission/Procedures** | X |  |  |
| **PROMIS Participation and Physical Function** | X | X |  |
| **PROMIS Pain Intensity and Pain Interference** | X | X |  |
| **VR-12** | X | X |  |
| **Complication & Adverse Event** | X | X |  |
| **Blood Draw** (*plasma*) | X | X | X |

**Table 2.**
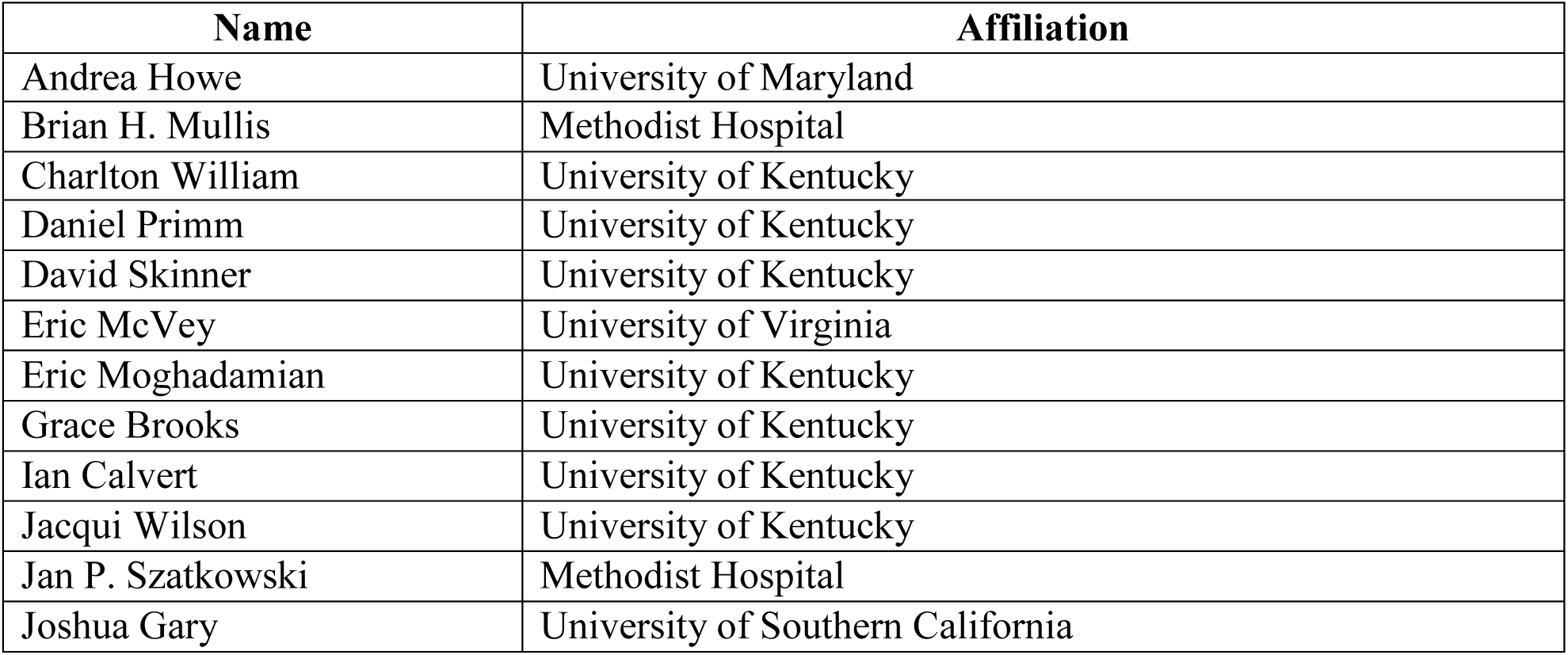
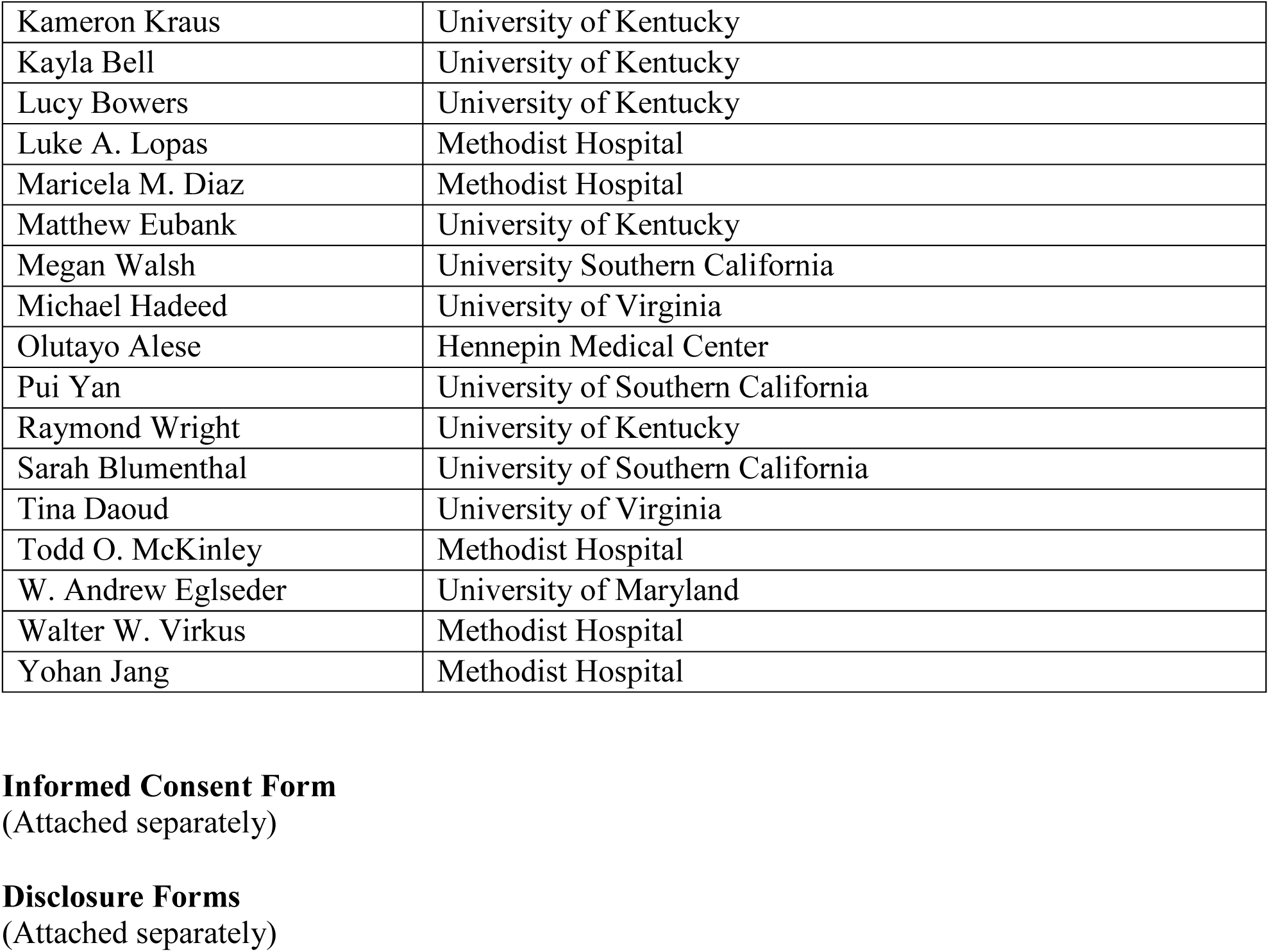
Consortium Authors and Affiliations.

